# Germline determinants of risk and molecular subtype in young-onset lung cancer

**DOI:** 10.64898/2026.06.30.26356693

**Authors:** Jaclyn LoPiccolo, Ryan L. Collins, Noah Fields, Carter Nakagawa, Kodi Taraszka, Xinan Wang, Li Su, Diane R. Koeller, Alison Levine Schwartz, Alicia Charleston Pollaci, Sarah M. Young, Victoria G. Williamson, Jose A. Avila, Emma Voligny, Tom Nguyen, Andy J. Pangilinan, Richard M. Erwin, Barbara J. Gitlitz, Silvia Novello, Geoffrey R. Oxnard, Ugonma N. Chukwueke, Priscilla K. Brastianos, Ayal A. Aizer, Mizuki Nishino Hatabu, Narjust Florez, Kevin M. Haigis, Eliezer M. Van Allen, Jorge J. Nieva, Judy E. Garber, David C. Christiani, Pasi A. Jänne, Alexander Gusev

## Abstract

Young-onset lung cancer is enriched for never-smoking and oncogene-driven tumors, yet its inherited genetic basis remains poorly defined. We performed germline whole-genome sequencing in 251 young-onset lung cancer cases (median age 37), which we jointly analyzed with never-smoking cases (n=196; median age 68) and cancer-free controls (n=1,883). We identified enrichments of rare deleterious coding variants across 55 cancer-related gene sets, including *EGFR/ERBB2* signaling and genes implicated by prior lung cancer GWAS. Exome-wide analyses of rare coding variants affirmed *TP53* as a penetrant lung cancer predisposition gene (odds ratio [OR]=36.1, p=1.02×10^-7^) and discovered two novel exome-wide significant tumor subtype-dependent associations: *IREB2* in cases with fusion-driven tumors (p=1.39×10^-6^) and *SMAD6* in fusion-negative tumors (p=2.05×10^-6^). Structural variants contributed distinct risk, with enrichment in constrained, lung-expressed genes (OR=5.79, p=5.8×10^-5^) and very large germline deletions being markedly enriched in cases with fusion-driven tumors. Polygenic risk scores for lung cancer were inversely correlated with rare variant burden, consistent with additive risk from rare and common variants. Collectively, these findings delineate a complex germline architecture underlying susceptibility and molecular subtype in young-onset lung cancer.

## Introduction

Lung cancer is the leading cause of cancer mortality worldwide, with a median age at diagnosis of ∼70 years. However, approximately 1,500-2,000 cases annually occur before age 45, many in individuals who have never smoked^1^. These young-onset tumors are enriched for distinct clinical and molecular features, including lung adenocarcinoma (LUAD) histology and targetable oncogenic fusions involving *ALK* and *ROS1*^2^. The early age-of-onset and diminished contribution from environmental risk in this population imply that germline genetics is a likely source of risk, offering a model to study inherited predisposition to oncogene-driven tumorigenesis in the absence of tobacco exposure. Prior genomic research in lung cancer has largely focused on somatic alterations and mutational signatures in tumors^3^. Most germline studies of lung cancer have focused on coding variation using gene panels or whole-exome sequencing^4,5^ and have identified rare pathogenic variants in established cancer predisposition genes, sometimes with evidence of germline-somatic interactions influencing tumor evolution^5-8^. However, few well-powered, genome-wide analyses of young-onset lung cancer cases have been performed to date^9^. Furthermore, targeted sequencing assays are inherently limited in their ability to capture structural, common, and noncoding variation. In contrast, whole-genome sequencing (WGS) enables a comprehensive assessment of the full spectrum of germline variation across mutational frequencies, contexts, and sizes^10,11^. Here, we generated the largest germline WGS cohort of young-onset lung cancer patients to date (n=251; median age 37) and jointly analyzed these data with 196 never-smoking LUAD cases and 1,883 adult controls^12,13^. By comprehensively evaluating rare, common, coding, noncoding, and structural variation in lung cancer predisposition and tumor evolution, our results demonstrate that young-onset lung cancer exhibits increased burdens of rare germline variants and polygenic risk, supporting a central role for inherited susceptibility in early-onset lung cancer biology.

## Results

### Clinicogenomic characterization of young-onset lung cancer patients

We first wanted to gain a clearer understanding of the distinct biology found in early-onset lung cancers by analyzing 24,774 LUAD samples from the AACR Project GENIE (v19.0)^14^. Consistent with prior reports^15,16^, we observed a strong association between young age and oncogenic fusions, including *ALK, ROS1*, and *RET*. Compared with patients diagnosed over age 45, those 45 and under had markedly higher odds of somatic fusion drivers (odds ratio [OR]=7.32, p=2.34×10^-64^), including *ALK* (OR=8.32, p=1.41×10^-42^), *ROS1* (OR=5.89, p=3.97×10^-14^), and *RET* (OR=3.55, p=1.60×10^-6^; **Figure 1, Supplementary Table 1**), with the odds of somatic *ALK* rearrangements decreased by 6.6% for each year of age at diagnosis (p=2.73×10^-65^), and odds of *KRAS* mutation increased by 1.5% per year (p=2.66×10^-28^)(**Supplementary Table 1**). Young patients also exhibited slightly higher odds of somatic *EGFR* (OR=1.20, p=2.87×10^-2^) and *HER2* mutations (OR=1.88, p=1.45×10^-3^). Conversely, drivers less frequent in younger patients included *KRAS* (OR=0.32, p=4.30×10^-35^), *MET* (OR=0.14, p=4.23×10^-6^), and *BRAF* (OR=0.36, p=1.52×10^-3^).

**Figure 1.**
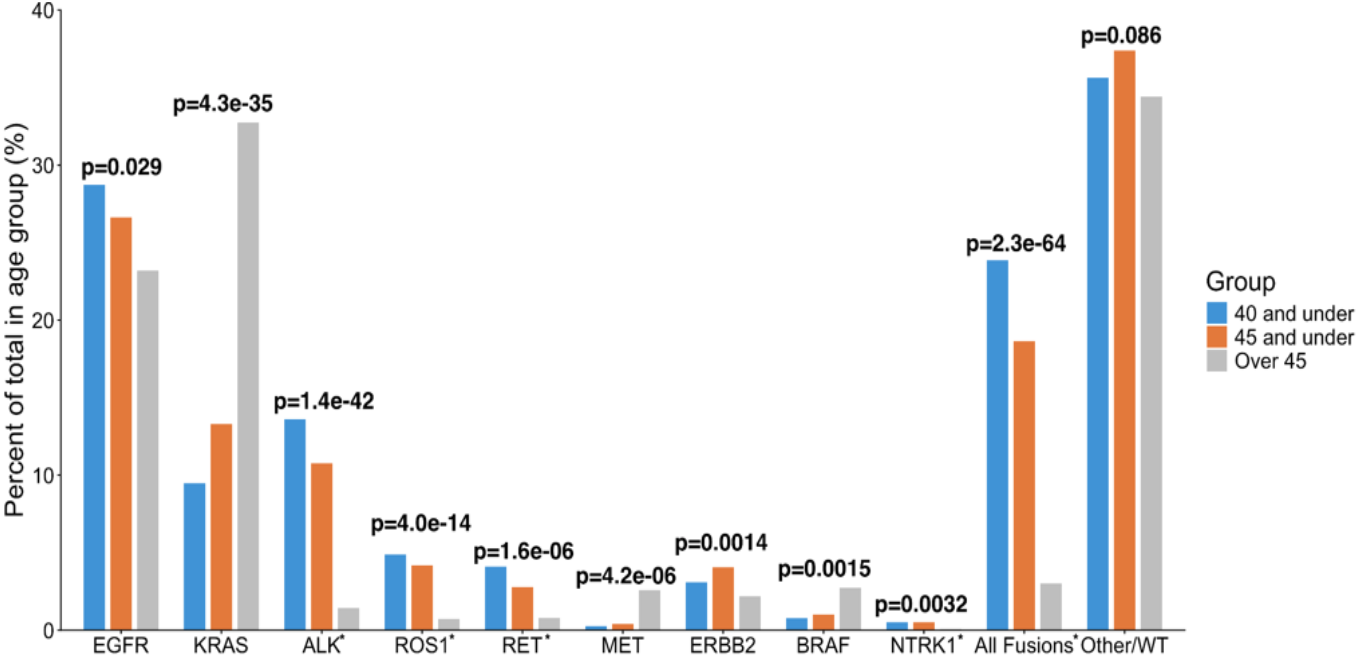
Association of age with oncogenic driver alterations in LUAD. Distribution of oncogenic driver alterations^37^ by age group in 24,774 lung adenocarcinoma (LUAD) cases from AACR Project GENIE (version 19.0). The proportion of tumors harboring specific driver alterations (defined here as OncoKB level 1 annotations) is shown for patients ≤40 years, ≤45 years, and >45 years at time of sequencing. Oncogenic fusions (*). WT/ unknown: negative for driver genes listed. Odds ratios (ORs), 95% coanfidence intervals, and P values for enrichment or depletion of each driver alteration among patients ≤45 versus >45 years, estimated using two-sided Fisher’s exact tests in **Supplemental Table 1**.

Motivated by the unique tumor biology of early-onset lung cancers, we assembled a cohort of 251 young-onset lung cancer patients treated at the Dana-Farber Cancer Institute between years 1997-2021 or recruited remotely from across the United States and Europe. This cohort size is equivalent to approximately 15% of young-onset non-small cell lung cancer (NSCLC) cases diagnosed annually in the United States^1^. All patients were diagnosed with NSCLC or small-cell lung cancer (SCLC) by age 45, and standard-of-care somatic testing for targetable driver alterations was performed at diagnosis. This cohort was predominantly female (60.6%), enriched for never-smoking history (73.9%), stage IV disease (76.1%), LUAD histology (90.8%), and presence of somatic targetable driver alterations (81.3% of entire cohort and 89.5% of LUAD; **Table 1)**. Among LUAD patients, somatic *EGFR* mutations were most common (39.0%), 69.7% of which were exon 19 deletions, followed by *ALK* fusions (23.3%; **Supplementary Table 2**). Overall, 30.7% of young LUAD cases harbored oncogenic fusions (e.g., *ALK, RET, ROS1*, and *NTRK*), representing a marked enrichment compared to 3.1% in older-onset LUAD from The Cancer Genome Atlas (TCGA; OR=15.8, Fisher’s exact test, p=5.35×10^-25^; median age 67 years). Somatic *EGFR* mutations were also enriched relative to TCGA (39.0% vs 10.4%; OR=5.49, p=1.3×10^-18^), whereas *KRAS* mutations were less frequent (10.1% vs 30.1%; OR=0.26, p=4.57×10^-9^; **Table 1**). Among stage IV LUAD in our cohort, female sex and presence of somatic fusions were associated with improved survival, exhibiting nearly two-fold improvement in overall survival (hazard ratio [HR]=0.51, 95% confidence interval [CI]=0.33–0.77, p=0.004) for fusion-positive cases versus fusion-negative, and HR=0.56 (95% CI=0.36–0.86, p=0.013) for *ALK*-rearranged LUAD versus non-ALK cases (**Supplementary Fig. 1**). These findings reinforce that early-onset NSCLC is composed of distinct molecular subsets associated with differential outcomes, warranting deeper characterization of germline and somatic factors.

**Table 1.**
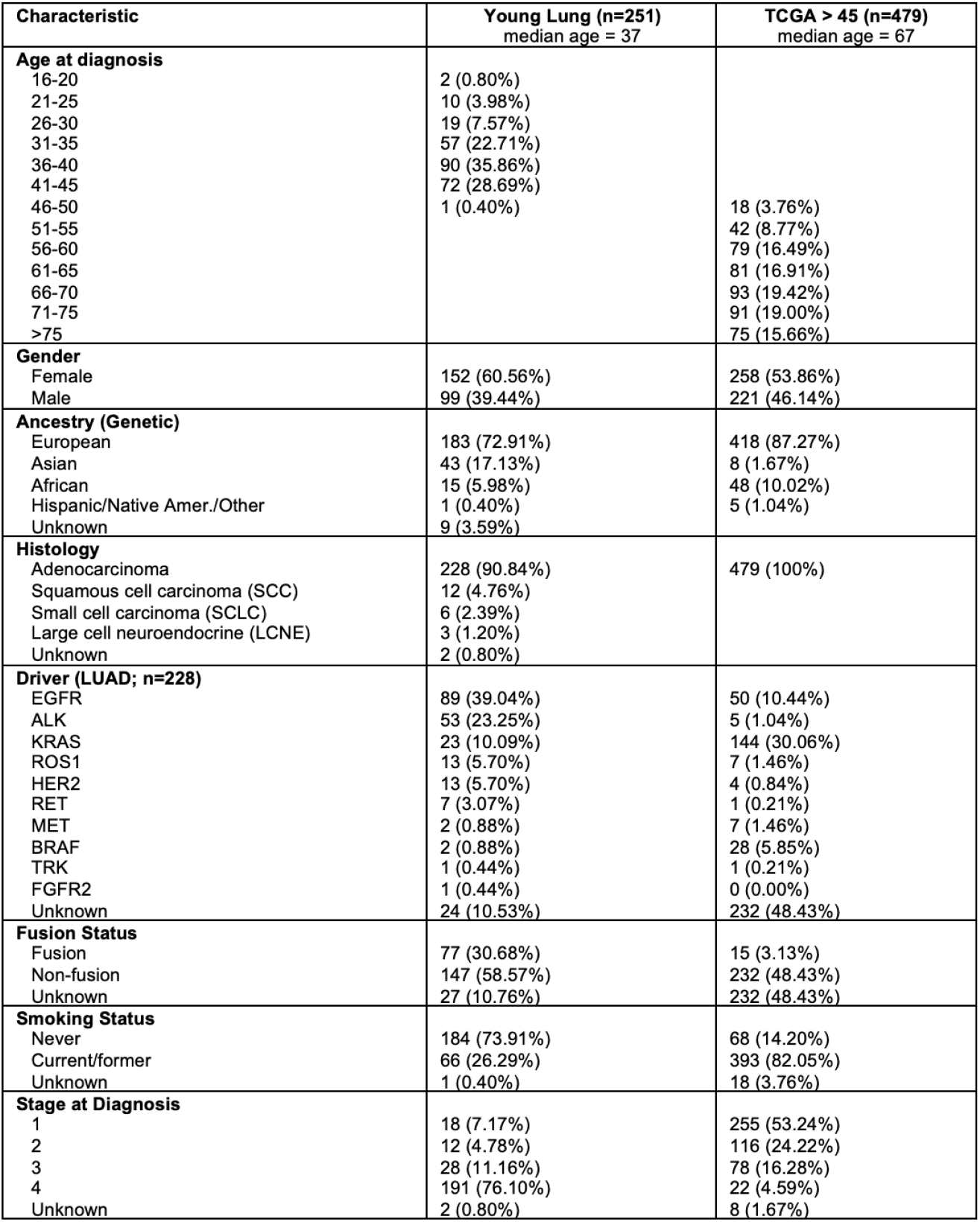
Clinical, genomic, and demographic characteristics of young-onset lung cancer and TCGA cohorts.

### The landscape of short and structural germline variation in young-onset lung cancer patients

We performed short-read WGS (median 43x coverage) from peripheral blood samples for all 251 patients, and jointly analyzed these genomes alongside two existing, technically comparable WGS datasets: (1) never-smoking LUAD from the Sherlock-Lung cohort (n=196)^12^ and (2) ancestry- and sex-matched adult controls from the Mt. Sinai BioMe Biobank (n=1,883)^13^. We detected and jointly genotyped single-nucleotide variants (SNVs), short insertions and deletions (indels; <50bp), and structural variants (SVs; ≥50 bp) in all 2,330 samples using two sister pipelines, GATK-HC and GATK-SV (**Figure 2a**)^17,18^. Together, these pipelines yielded high-quality, jointly genotyped SNV/indel data for all 2,330 samples; following SV-specific *post hoc* quality control and outlier sample exclusion, high-quality germline SV data was also retained for 2,166 samples. Across these 2,166 samples with complete SNV, indel, and SV data, we detected a total of 76M SNVs, 14M indels, and 287k SVs carried in at least one genome (**Figure 2b**), which were broadly concordant with gold-standard reference datasets from the Genome Aggregation Database (gnomAD) v4.1 ^19^ (*i*.*e*., 96.8% of all variants with allele frequency [AF]>1% were also reported in gnomAD; **Supplementary Fig. 2**). Principal component analyses of common variants confirmed that this cohort was largely genetically similar to European ancestral populations (85.3%), with East Asian ancestry representing the second most abundant population (7.2%; **Supplementary Fig. 3**). Importantly, WGS enabled us to capture an average of 9,433 SVs per genome across seven mutational subclasses with no meaningful genome-wide differences between cases and controls (**Supplementary Fig. 4**)^11^. Most SVs detected in this cohort were small (median size=92bp; **Figure 2c**) and rare (86% had AF<1%; **Figure 2d**), and therefore comprised a tranche of germline variation that has been underexplored in early-onset lung cancer and cancer predisposition more broadly.

**Figure 2.**
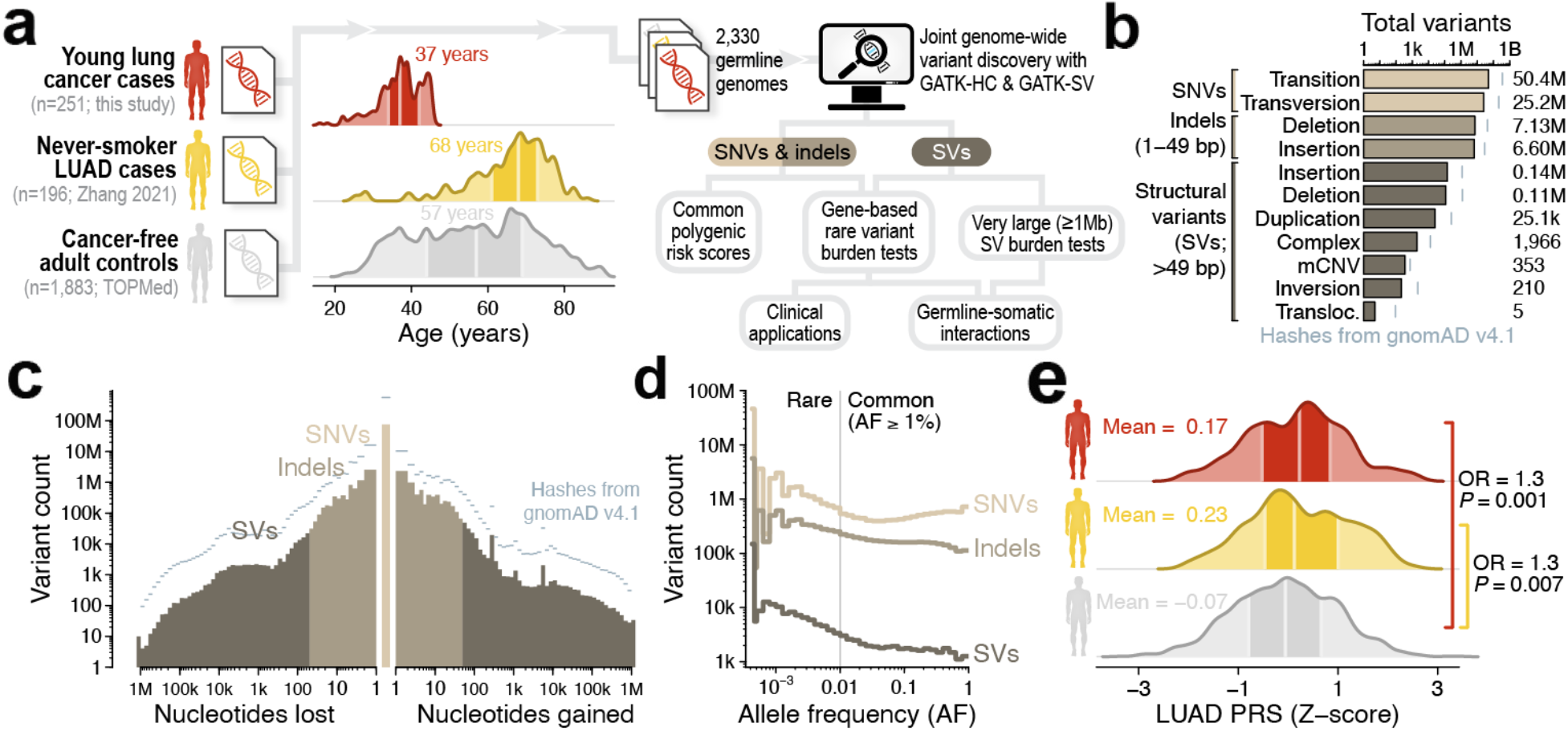
Characterizing germline variation in lung cancer cases and cancer-free controls. (**a**) We generated short-read germline WGS for 251 young-onset lung cancer cases and jointly analyzed these data alongside published WGS from 196 never-smoker LUAD cases and 1,883 cancer-free adult controls ^12^. (**b**) We detected a total of 89.6M distinct germline variants carried in at least one genome in our cohort. Grey hashes depict variant counts from the Genome Aggregation Database v4.1 for comparison ^13^. (**c**) Our pipelines captured the full range of variant sizes. (**d**) Most (84.1%) variants detected in this cohort were rare (AF<1%). (**e**) Polygenic risk scores (PRS) developed in prior LUAD case-control studies were associated with modest risk for both young lung and never-smoker LUAD compared to controls (logistic regression of European ancestry cases & controls adjusted for sex and five European-specific principal components).

### Polygenic risk for lung adenocarcinoma generalizes to early-onset and never-smoking populations

We first investigated whether polygenic risk scores (PRS) derived from recent lung cancer GWAS were associated with young-onset lung cancer. We calculated lung cancer and LUAD-specific PRS for all 2,330 individuals based on a recent cross-ancestry GWAS meta-analysis of a wide range of lung cancer populations^20^, most of which would be expected to have smoking-related disease and average onset age (∼70 years). In European individuals, LUAD PRS was significantly associated with both young-onset lung cancer (p=0.001, OR=1.31 per standard deviation increase in PRS, 95% CI=1.11-1.53) and never-smoker LUAD from the Sherlock-Lung cohort (p=0.007, OR=1.31, 95% CI=1.08-1.60; **Figure 2e, Supplementary Table 3**). Controlling for participant age in the young-onset analysis by restricting to controls under 45 years old did not alter the association between LUAD PRS and early-onset lung cancer (OR=1.37; 95% CI=1.13-1.65; p=0.001; n=604 cases/controls). Given the comparable PRS associations observed for young-onset and older, never-smoking lung cancer cases, we conclude that elevated polygenic risk does contribute to young-onset patients but does not appear to be the primary explanation for early disease onset. Additionally, we did not identify associations between LUAD PRS and survival, which may suggest that the polygenic risk associated with lung cancer initiation exerts lesser influence on lung cancer disease course, as has been recently demonstrated for other common, complex diseases^21,22^.

### Young lung cancer cases are enriched for rare damaging germline variants in cancer-associated genes

We turned to rare variants as another potential source of risk for early-onset lung cancer. We first tested whether young-onset lung cancer cases were enriched for rare (AF<1% in gnomAD and singletons or doubletons in our cohort) coding variants in a curated set of 154 gene sets and pathways implicated in pro-survival signaling, lung cancer/biology, or cancer genetics (**Supplementary Table 4**). We next classified rare coding SNVs and indels into four tiers based on predicted functional impact: (1) pathogenic/likely pathogenic (P/LP) variants in ClinVar (“CV”); (2) ClinVar P/LP or Ensembl Variant Effect Predictor (VEP)^23^ high-impact loss-of-function variants (“CV-HIGH”); (3) ClinVar P/LP, VEP high-impact, or missense variants with rare exome variant ensemble learner (REVEL)^24^ score >0.5 (“CV-HIGH-REVEL-0.5”); and (4) all VEP high- or moderate-impact variants (“HIGH-MOD”). Finally, we tested each gene set and variant consequence tier for association with case status in individuals of European ancestry from the young-onset cohort (n=171), the Sherlock-Lung LUAD cohort (n=182), or the two cohorts combined (n=353), as compared to controls (n=1,606), while adjusting for sex and five European-specific ancestry components.

We identified significant enrichments of rare, gene-disruptive coding variants in 23/154 gene sets among individuals with young-onset lung cancer compared to non-cancer controls surpassing Benjamini-Hochberg false discovery rate (FDR) <0.05 (**Figure 3a**; **Supplementary Table 5**), with an additional 22/154 at unadjusted nominal significance (p<0.05). Key findings included enrichments of clinically interpretable (“CV”) rare variants in *TP53* (OR=32.9, p=4.02×10^-4^, FDR=6.00×10^-2^), loss-of-function-intolerant cancer predisposition genes (OR=6.42, p=5.48×10^-4^, FDR=1.92×10^-2^), tumor suppressors (OR=6.64, p=1.03×10^-4^, FDR=1.47×10^-2^), and NSCLC signaling pathways (OR=8.34, p=4.87×10^-4^, FDR=1.92×10^-2^). Strong effects from CV-tier variants were also observed in the early-onset cohort for other canonical cancer pathways at nominal significance, such as EGFR-interacting genes (OR = 13.79, p=3.45×10^-2^). These rare variant effects were less pronounced in never-smoker LUAD cases (**Figure 3b-c**), with just 16/154 gene sets exhibiting significant enrichments (FDR<0.1) of rare variants specific to never-smoker cases.

**Figure 3.**
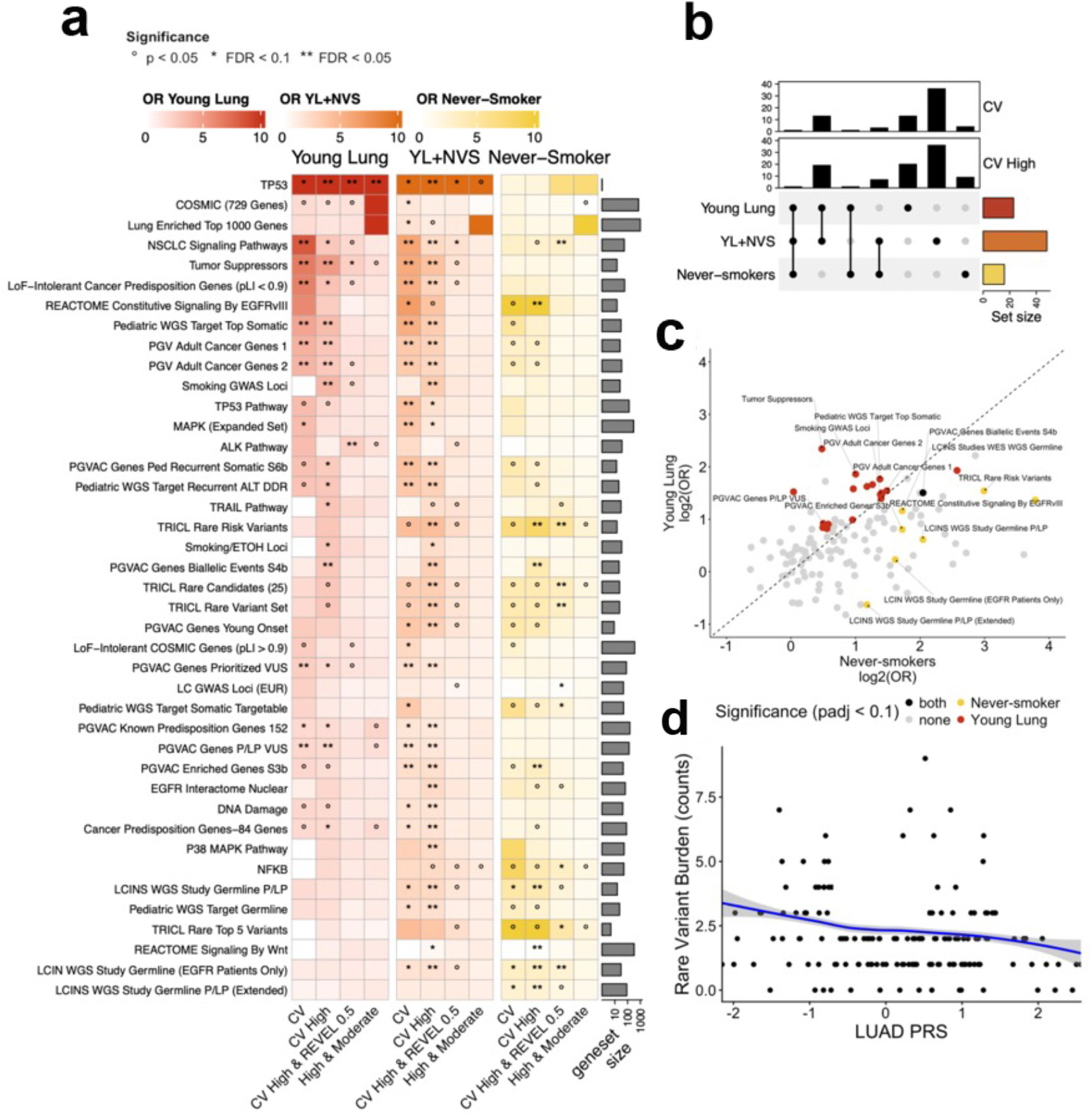
Enrichment of rare coding variants in cancer gene sets. (**a**) Summary heatmap of odds ratios from geneset burden tests for four tiers of rare coding variants (columns) and select cancer-relevant gene sets (rows). Significant gene sets from any category are included (p<0.01, q<0.1). Significance is indicated as follows: **q<0.05, *q<0.1, °p<0.05. (**b**) UpSet plot of significant genesets (q<0.1) between cohorts for CV and CV-HIGH tiers. (**c**) Comparison of rare CV-HIGH variant burden odds-ratios in young-lung and never-smoker cohorts. Points are colored by significance (q<0.1), with highly significant (q<0.05) genesets labeled. (**d**) Scaled LUAD PRS vs. rare CV-HIGH variant burden in enriched gene set genes for the young lung cohort. The trendline shown is derived from Poisson model predictions on sample data.

Given recent reports of PRS correlating with rates of rare P/LP germline variants in other common adult cancers^25^, we next assessed whether LUAD PRS was associated with rates of rare coding variants among lung cancer cases in our cohort. Indeed, in young-onset LUAD patients LUAD PRS was significantly negatively correlated by Poisson regression with rare CV variant burden in young-onset enriched genes from gene set analysis (β=-0.429, p=2.16×10^-3^; **Figure 3d; Supplementary Table 6**). In contrast, rare, synonymous variants in the same gene sets did not show any correlation with LUAD PRS (β=-0.041, p=0.198). These findings suggest that the genetic architecture of early-onset lung cancer likely follows a liability threshold model wherein either many weak-effect common risk SNPs or a small number of strong-effect rare coding variants is sufficient to promote lung tumorigenesis unusually early in life.

### Exome-wide discovery of new candidate lung cancer predisposition genes

We hypothesized that the rare variant enrichments we observed in curated gene sets were driven by specific variants concentrated in individual predisposition genes, including as-of-yet undiscovered predisposition genes. We therefore performed exome-wide gene-based association testing of 18,544 autosomal protein-coding genes to identify lung cancer predisposition genes. For each gene, we compared the rare (AF<0.1%) coding variants across the four annotation tiers used in our burden analyses between European ancestry cases and controls using logistic mixed models accounting for genetic relatedness and sex^26^. We performed five exome-wide gene discovery analyses, one for each case set including: (1) all cases, (2) young-onset cohort, (3) never-smoking cohort, (4) patients with fusion-driven cancers (n=80), and (5) patients with non-fusion-driven cancers (n=253).

Across these analyses, five genes reached strict exome-wide significance (p<2.69×10^-6^): *TP53, SMAD6, IREB2*, and *ZNF678* with tier 3 (“CV-HIGH-REVEL-0.5”) variants, and *MYH9* with tier 2 (“CV-HIGH”) variants (**Figure 4a-c; Supplementary Table 7**). Only *TP53* reached significance when comparing all lung cancer cases to controls: rare damaging *TP53* variants were strongly associated with overall lung cancer risk (8/353 cases vs 3/1,606 controls; OR=36.1; 95%; p=1.02×10^-7^). Carriers were usually younger at diagnosis (median 38 years)^27^, and variants clustered within the DNA-binding domain, a known hotspot for pathogenic *TP53* mutations underlying Li-Fraumeni Syndrome (**Figure 4d**)^28^. Beyond *TP53*, the other four genes achieved exome-wide significance only for specific subsets of cases. *IREB2* was associated exclusively with fusion-positive tumors (4/80 cases vs 1/1,606 controls; p=1.39×10^-6^), whereas *SMAD6* was significant only in patients with non-fusion drivers (4/253 cases vs 0/1,606 controls; p=2.05×10^-6^). We performed sensitivity analyses for *IREB2* and *SMAD6* by comparing variant carrier frequencies between fusion- and non-fusion-driven cases, finding that rare damaging variants in *IREB2* were significantly enriched among fusion-positive vs. -negative cases (p=0.01). No significant association within cases was seen for *SMAD6*, likely due to small sample sizes. The final two significant genes were associated with lung cancer risk exclusively in never-smokers: *MYH9* (p=6.23×10^-7^), which has been implicated in lung function by prior GWAS and reported as a low-frequency somatic mutational driver in 2-3% of NSCLC tumors ^29-31^, and *ZNF678* (p=2.58×10^-9^), which has not yet been robustly implicated in any cancer to our knowledge.

**Figure 4.**
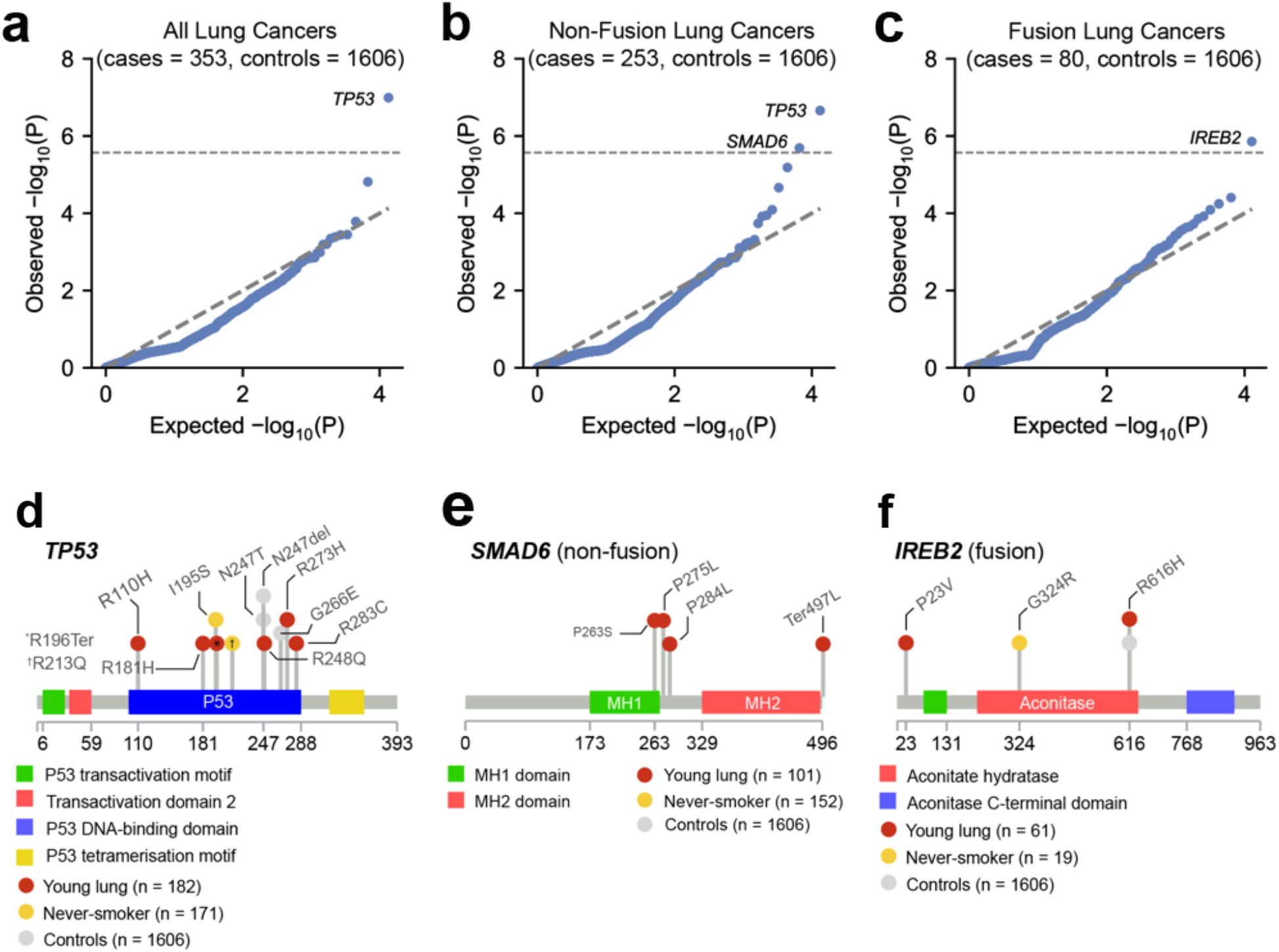
Exome-wide discovery of lung cancer predisposition genes. (**a-c**) Quantile-quantile plots for exome-wide discovery of candidate predisposition genes for (**a**) all lung cancer cases, (**b**) cases with non-fusion driven tumors, and (**c**) cases with fusion-driven tumors. Across all exome-wide discovery analyses, we identified three genes exceeding strict Bonferroni significance (p<2.69×10-6): (**d**) *TP53* in all cases and controls, (**e**) *SMAD6* in non-fusion driven cases, and (**f**) *IREB2* in fusion-driven cases.

Several other established cancer predisposition genes demonstrated nominally significant enrichments in cases vs. controls, including *ATM* (never-smokers; p=1.07×10^-4^; median age=66). Given that both *ATM* and *TP53* are central to DNA double-stranded break sensing, we explored the somatic driver status of germline *ATM* and *TP53* carriers, finding that all carriers developed non-fusion-driven lung cancers. While this trend towards non-fusion-driven tumors was not individually significant for either *ATM* or *TP53* alone, collectively these genes produced a nominally significant association within cases favoring non-fusion-driven lung tumors (14/14 carriers vs. 239/319 non-carriers; p=0.026, two-sided Fisher’s exact test).

### Rare germline structural variants predispose to early-onset lung cancer

SVs are an understudied source of variation present in every germline genome that can have profound influences on disease risk^11^. In light of our previous findings implicating rare, protein-coding SNVs/indels in lung cancer risk, we similarly hypothesized that rare germline SVs might confer lung cancer risk by disrupting specific sets of genes. We tested this hypothesis by performing collapsing burden tests of rare loss-of-function SVs in each of the gene sets between European cases and controls as previously for short variants. Although germline SVs were an order of magnitude sparser than germline SNV/indels, we nonetheless identified three gene sets significantly enriched (FDR q<0.05) in rare loss-of-function SVs (**Supplementary Table 8**). All three of these gene sets corresponded to mutationally constrained genes that were highly expressed in adult lung tissue^32,33^. For example, mutationally constrained genes among the top 1,000 most highly lung-expressed genes were disrupted by rare SVs in 5.5% (17/309) of cases vs. 1.6% (22/1,411) of controls (OR=4.64, p=6.7×10^-5^). This signal was most pronounced in young lung patients (OR=5.79, p=5.8×10^-5^; **Figure 5a**) and was not observed in never-smoking cases from the Sherlock-Lung cohort (OR=1.71, p=0.32; **Figure 5b**).

**Figure 5.**
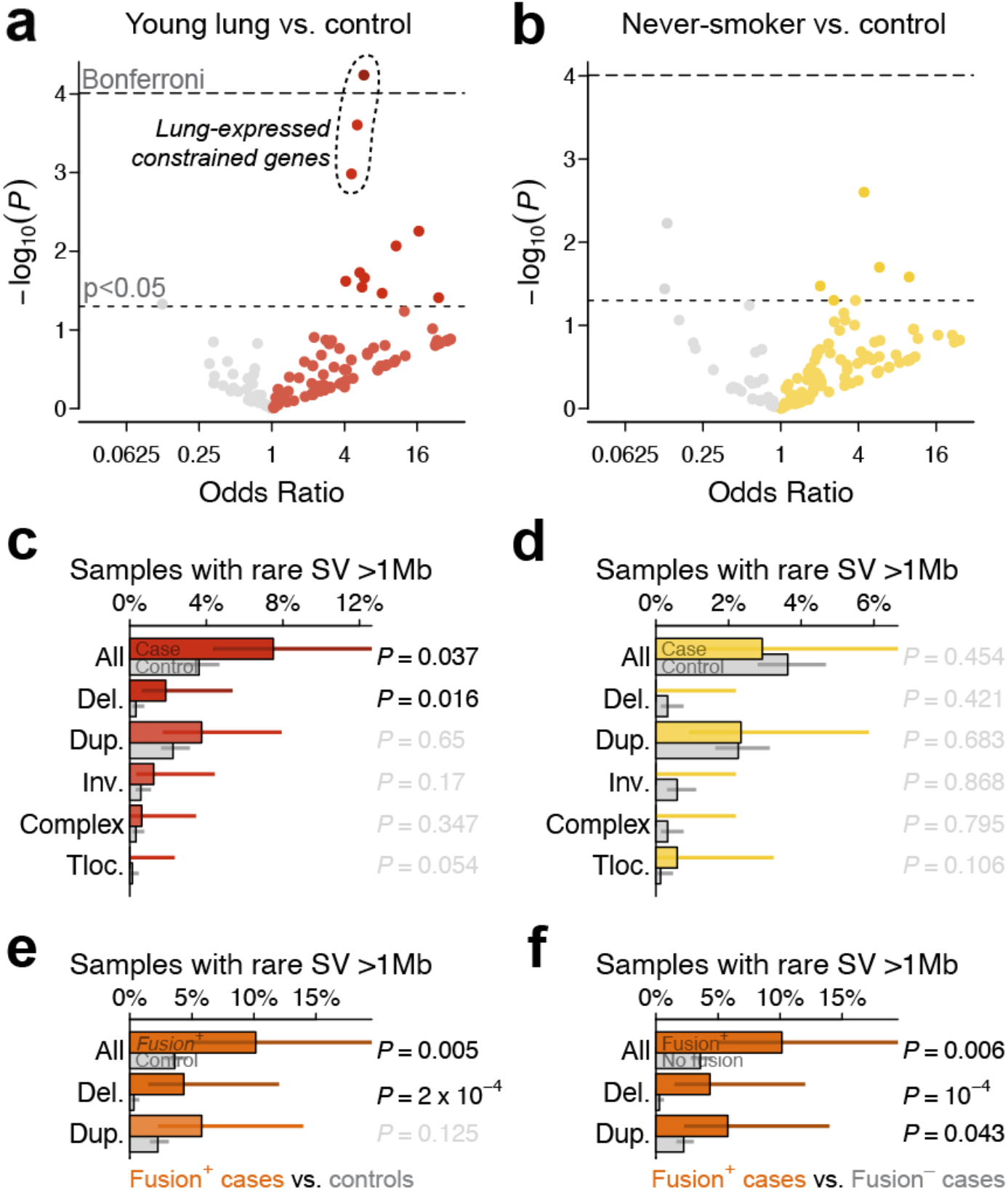
Large and coding rare SVs are associated with early-onset and fusion-driven lung cancer. (**a-b**) Rare loss-of-function SV burdens in 154 gene sets between (**a**) young lung cases or (**b**) never-smoker LUAD cases from the Sherlock-Lung cohort versus controls of European ancestry. The three outlined tests in (**a**) surpass Benjamini-Hochberg FDR q<0.05. (**c-d**) The rate of exceptionally large (>1Mb) rare germline SVs was significantly higher in (**c**) young lung cases versus controls, but not in (**d**) never-smoker LUAD cases from Sherlock-Lung. (**e-f**) The excess of large, rare germline SVs was most pronounced in lung cancer patients with fusion-driven tumors and was significant when comparing fusion-positive cases to either (**e**) cancer-free controls or (**f**) lung cancer patients with fusion-negative tumors.

We further hypothesized that germline SVs might also act through mechanisms beyond direct gene disruption, such as by sensitizing to somatic genome instability, as has been recently proposed in pediatric solid tumors^34,35^. We performed case-control association tests for genome-wide SV burdens of large (≥1Mb), rare (AF<1%) SVs, finding a nominally elevated rate in European-ancestry young-onset lung cancer cases versus controls after adjusting for European-specific principal components and sex (12/160 cases vs. 56/1,545 controls; OR=2.5, 95% CI=1.1-5.8, p=0.037; **Figure 5c**), but no such enrichment in never-smoker cases from the Sherlock-Lung cohort (p=0.45; **Figure 5d**). Stratifying this genome-wide burden by SV class revealed that this enrichment was most pronounced for large deletion SVs (N=8 SVs; 3/160 young lung cases vs. 5/1,545 controls; OR=11.3, 95% CI=1.6-81.5, p=0.02). We scrutinized the raw sequencing evidence for each of these variants, finding unambiguous support consistent with germline origin for all large, rare deletions (**Supplementary Figure 5**).

Curiously, none of these large, rare deletions in cases could be explained by overlap with any known cancer genes^36^. In fact, the only overlap between a large germline deletion and COSMIC gene in our dataset was identified in a control, who carried a 1.7Mb deletion predicted to cause loss-of-function of *RGPD3*. Upon scrutinizing these three large deletions in cases, all were carried by LUAD patients with tumors driven by *ALK* fusions, a significant enrichment when comparing fusion-positive cases either to controls (OR=17.5 p=1.8×10^-4^; **Figure 5e**) or to fusion-negative cases (OR=18.2, p=1.3× 10^-4^; **Figure 5f**). Notably, none of these deletions occurred on chromosome 2, which contains the *ALK* locus, arguing against a direct structural effect on *ALK* as the explanation for their enrichment among *ALK*-rearranged tumors. When taken together with the *SMAD6* and *IREB2* rare variant associations we discovered above, these SV findings further support a model in which specific germline factors shape tumor evolution by predisposing carriers to specific somatic driver events.

### Rare noncoding germline variation in early-onset lung cancer risk

Having established the contribution of rare coding and structural germline variants to early-onset lung cancer risk, we next evaluated rare noncoding germline variant carrier status using candidate regulatory elements defined in lung tissue. Two candidate gene sets were evaluated: (1) 27 tumor suppressor genes and (2) 11 genes comprising known LUAD drivers (*EGFR, ALK, ERBB2, KRAS, BRAF, MET, RET, ROS1*)^37^ or with exome-wide significant associations with early-onset patients in our study (*TP53, SMAD6, IREB2*). Case-control analyses in the young-onset cohort and the combined young-onset and never-smoking cohort identified no significant gene- or gene-set associations (**Supplementary Table 9**). Although larger studies are needed, these findings suggest rare noncoding variants in candidate *cis*-regulatory elements contribute less risk than rare coding or structural variation.

### Clinical response to germline-directed therapy in a young lung cancer patient

Treatment selection in advanced NSCLC is typically guided by somatic genotyping, with germline variants rarely informing therapy. However, one stage IV LUAD patient in our young-onset cohort, with a pathogenic germline *BRCA2* variant, tumor loss of heterozygosity, and no alternative somatic driver experienced an exceptional response to germline-directed treatment. Given sensitivity of *BRCA2*-deficient tumors to DNA-damaging agents and PARP inhibition, the patient received platinum-based chemotherapy for approximately 55 months followed by PARP inhibition with niraparib, with survival exceeding six years compared with benchmark outcomes for stage IV NSCLC (median OS 7–10 months with chemotherapy alone, 5-year survival with distant metastases ∼12%, and 5-year survival with brain metastases approximately 4-6%). Serial brain MRI demonstrated radiographic response of intracranial lesions (**Figure 6a-b**), with no evidence of systemic disease since starting chemotherapy (**Figure 6c**). This case underscores the potential clinical utility of germline findings in guiding treatment.

**Figure 6.**
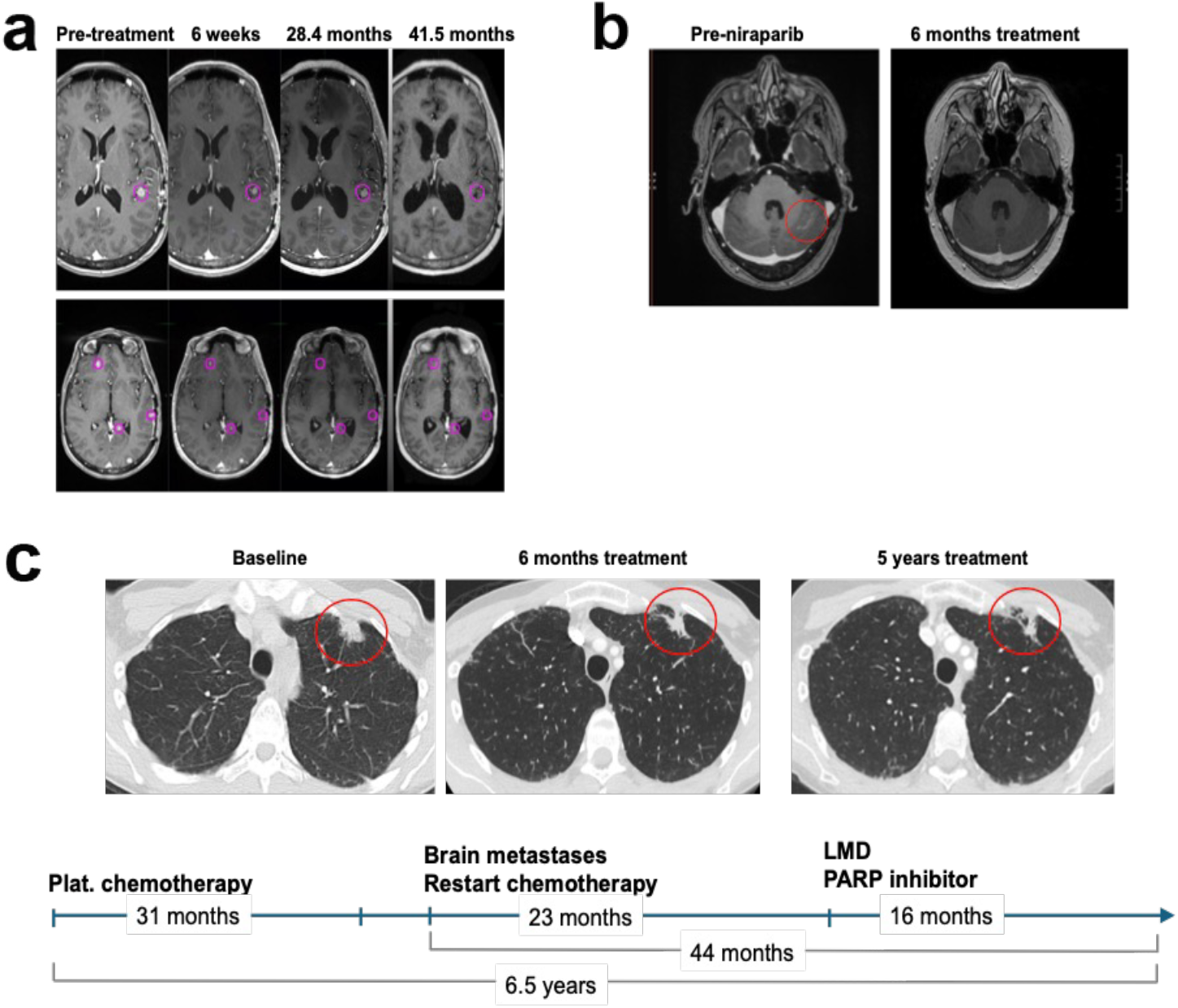
Response to germline-directed therapy in a young-onset lung cancer case with a pathogenic germline *BRCA2* variant. (**a**) Serial contrast-enhanced brain MRI demonstrating radiographic response and eventual resolution of representative intracranial metastases following platinum-based chemotherapy and subsequent PARP inhibition; these lesions did not receive radiation. Representative lesions are circled. (**b**) Additional posterior fossa metastasis demonstrating interval resolution after treatment. (**c**) Serial chest CT imaging showing sustained control of the primary left upper lobe lung lesion without evidence of systemic progression during therapy. Timeline depicts treatment course, including approximately 55 months of platinum-based chemotherapy followed by niraparib maintenance, with ongoing survival exceeding 6.5 years from treatment initiation. The patient harbored a pathogenic germline *BRCA2* variant with tumor loss of heterozygosity and no alternative canonical oncogenic driver alteration.

## Discussion

In this study, we performed a comprehensive analysis of germline risk factors in early-onset lung cancer by integrating germline WGS with clinicogenomic annotations. This cohort is notable for both its scale and depth, reflecting over two decades of recruitment with clinical, germline, and somatic data. Moreover, by jointly analyzing young-onset lung cancer cases alongside never-smoking lung cancer patients of typical age from the Sherlock-Lung study^12^ as well as cancer-free controls, we identified multiple forms of germline variation associated specifically with early-onset disease and not with lung cancers found in older patients with a similar lack of tobacco smoke exposure. Prior pioneering studies from the Sherlock-Lung cohort^3,12^ provided foundational genome-wide assessments of the somatic landscape of LUAD in never-smoking patients, but the extent to which these patterns also applied to early-onset LUAD remained uncertain. Building upon the discoveries of Sherlock-Lung, we leveraged our new germline WGS data for young-onset lung cancer patients within a formal case-control framework to perform a systematic analysis of germline susceptibility across the full spectrum of genomic variation, including rare, common, structural, and noncoding variants, extending prior observations of germline variation in LCINS.

A central finding of this study is the robust association between damaging rare variants in *TP53* and early-onset lung cancer. *TP53* is the established cause of Li-Fraumeni syndrome (LFS); prior LFS-focused studies have demonstrated increased incidence of lung cancer at younger ages among carriers^38^, but these studies primarily described individuals with known LFS and did not establish whether rare damaging *TP53* variants were enriched among young lung cancer patients relative to cancer-free controls. Using germline WGS within a case-control framework and rigorous exome-wide association testing, we demonstrate that *TP53* PGVs are unambiguously enriched in young-onset lung cancer. We did not observe a similar enrichment in older-onset never-smoking cases, supporting a uniquely early-onset lung cancer predisposition in germline *TP53* carriers. Notably, many *TP53* carriers in this cohort were probands without prior family history of lung cancer, and only one of the eight *TP53* PGV carriers originated from a previously recognized LFS family. Most carriers therefore would not have been identifiable on the basis of a known familial LFS diagnosis alone, highlighting the potential value of germline testing in young-onset lung cancer patients. The identification of these individuals and their families has immediate implications for cascade testing and implementation of established LFS surveillance protocols.

Our gene association analyses also nominated *SMAD6* and *IREB2* as candidate germline susceptibility genes with potential molecular subtype-specific relevance: their tumor subtype-specific enrichment patterns may imply that distinct germline pathways predispose to different molecular subtypes of lung cancer. *SMAD6*, an inhibitor of BMP and TGF-β signaling, implicates dysregulated developmental and growth factor signaling in non-fusion-driven tumorigenesis, potentially modifying susceptibility in tumors characterized by MAPK pathway activation (*e*.*g*., activating *EGFR* or *KRAS* mutations). In contrast, *IREB2*, which regulates cellular iron homeostasis, points to a role for metabolic and oxidative stress pathways in shaping susceptibility to fusion-driven disease. Intriguingly, *IREB2* has been associated with lung cancer in multiple prior GWAS^39-41^, whereas rare coding variants in *IREB2* have been associated in the UK Biobank with both bronchitis (p=1.1×10^-4^; PheWAS rank #4 of 4,529) and whooping cough (p=9.6×10^-4^; #12 of 4,529)^42^. Together, these findings support a model in which germline variation impacting core cellular pathways influences not only cancer risk but also the frequency of specific oncogenic drivers.

Our results also provide a more complete view of the total genetic architecture of early-onset lung cancer. Specifically, we show that PRS, derived from population-based studies largely enriched for smoking-related disease, remain significantly associated with risk in this population. However, the inverse correlation observed between PRS and rare pathogenic variant burden supports a liability threshold model wherein rare, high-penetrance variants and common, low-effect alleles contribute independently to disease risk. This suggests that polygenic background contributes to lung cancer susceptibility even in the absence of tobacco exposure, while reinforcing the role of rare variation in early-onset disease.

A major strength of WGS is the ability to interrogate SVs, a class of genetic variation largely inaccessible to lower-resolution technologies like exome sequencing. By applying specialized pipelines, we identified rare germline SVs as a significant contributor to early-onset lung cancer risk. Consistent with expectations set by prior population sequencing studies, the genome-wide burden of all germline SVs *en masse* did not differ between cases and controls. Instead, we discovered significant enrichments of rare loss-of-function SVs in mutationally constrained, lung-expressed genes, as well as an increased burden of large (>1 Mb) rare deletions among young cases. Strikingly, these large germline deletions were exclusively observed in patients with *ALK* fusion-driven tumors, suggesting a previously unrecognized link between germline SVs and somatic genome stability. These findings highlight the importance of considering germline SVs in cancer predisposition and demonstrate the added value of WGS in uncovering mechanisms that would otherwise remain undetected.

Our study has several limitations. First, although our study reports the largest WGS cohort of early-onset lung cancer assembled to date, our sample size is still modest relative to genome-wide discovery efforts in other cancers. This limitation is magnified for molecular subtype-specific analyses with even smaller sample sizes, reducing power to detect subtype-specific associations and increasing uncertainty around effect estimates. As a result, findings in these subgroup analyses – such as the associations observed for *IREB2* in fusion-driven tumors and the enrichment of large germline SVs among *ALK*-positive cases – will require validation in larger, independent cohorts. Second, our analysis of regulatory regions was intentionally restricted to a focused set of loci with strong prior likelihood for relevance in lung cancer. This targeted approach reflects both the current limitations in functional annotation of the noncoding genome and the modest sample size available for rare variant analyses. As a result, we may have missed important regulatory variation outside of these regions. Larger cohorts and improved annotation frameworks will be required to enable unbiased discovery of noncoding germline risk factors. Third, our cohort was composed predominantly of individuals of European ancestry, reflecting the current necessity of ancestry-matched genetic analyses but limiting generalizability to underrepresented populations, particularly those with higher rates of *EGFR*-mutant lung cancer, including Asian and Hispanic/Latinx individuals.

Despite these limitations, our findings have important implications. First, they support the consideration of germline testing in young or never-smoking lung cancer patients, where the likelihood of identifying clinically actionable variants may be higher than previously appreciated. Identification of high-penetrance variants in genes like *TP53* can inform surveillance strategies for patients and family members. Second, our results highlight the potential for germline variation to inform therapeutic decision-making. The observed durable response to platinum-based chemotherapy and PARP inhibition in a patient with a germline *BRCA2* mutation underscores the relevance of HR deficiency in a subset of lung cancers and suggests that germline-directed treatment strategies may be clinically relevant in this disease. Finally, these findings raise the possibility that integrated germline risk models (including rare variants, SVs, and polygenic risk) could inform future screening strategies, particularly for individuals currently excluded from guideline-based screening due to age and lack of smoking history.

## Methods

### Ethics approval and consent to participate

Written informed consent for comprehensive genetic analysis of germline samples was obtained from participants in the original studies, which were approved by the Dana-Farber Cancer Institute institutional review board. Secondary genomic analyses conducted for this study were approved under DFCI-IRB protocols 02-180, 14-183, and 18-411. This research adheres to the principles outlined in the Declaration of Helsinki. All whole-genome sequencing data generated in this study will soon be deposited to the database of Genotypes and Phenotypes (dbGaP).

### Sample selection

Young-onset lung cancer patients were defined as onset of histologically confirmed non-small cell or small cell lung cancer at age 45 or younger, which in total included 251 patients. These patients were recruited through from three sub-studies: 1) DFLC: retrospectively identified patients who received care at the Dana-Farber Cancer Institute between years 1997-2018 (DF #02-180) (n=83); 2) GOYLC: patients from prior prospective study Genomics of Young Lung Cancer ^2^(DF #14-183) (n=108), and 3) PROCA: patients prospectively enrolled in Profile And Cancer gene Testing for Individual Evaluation; PROACTIVE (DF #18-411) (n=56). Peripheral blood samples were available for all participants, with fresh peripheral blood for PROCA and frozen for DFLC and GOYLC. Tumor driver mutations genotype were determined by clinical single or multi-gene panel sequencing; 37.5% (n=94) had somatic panel-based NGS using internal DFCI NGS assay Oncopanel. For germline WGS, genomic DNA was extracted from fresh or frozen peripheral blood using a standard nucleic acid isolation protocol by the Genomics Platform at the Broad Institute of M.I.T. and Harvard. Short-read WGS was performed on genomic DNA samples for all participants to a minimum of 30x coverage on an Illumina Novaseq instrument at the Broad Institute. Samples passing quality control metrics were aligned to the human reference genome (hg38).

### Comparison cohorts

We jointly analyzed the young-lung cancer cohort recruited and sequenced in this study alongside two existing published germline WGS datasets as comparison cohorts. First, we accessed germline WGS data from the dbGaP study “Integrative Analysis of Lung Adenocarcinoma in Never Smokers” phs001697.v1.p1 ^12^, which performed germline WGS on 232 patients with lung cancer without a history of smoking from Institut universitaire de cardiologie et de pneumologie de Québec - Université Laval (IUCPQ-UL) (n=125), Université Côte d’Azur (Nice) (n=47), the Environment And Genetics in Lung cancer Etiology (EAGLE) study (n=27), Yale University (n=22), and Moffitt Cancer Center (n=11). We accessed germline WGS data for all 232 samples and realigned this data to the hg38 reference genome using the same methods as for our young-onset lung cancer cohort. Second, we selected 1,883 cancer-free controls from Mt. Sinai BioMe Biobank ^13^, matching to cases on reported ancestry and sex in a 4:1 ratio. Given our study’s emphasis on early-onset lung cancer, we accounted for age of controls using a stratified approach by selecting four equal-sized sets of controls to achieve an overall 4:1 ratio of controls to cases: one set of controls was strictly matched to case ages, one set included older controls (>65 years old) who we reasoned were at or beyond the average age-of-onset for lung cancer (∼68 years old), and two sets of controls were selected at random to reflect the general adult population included in the BioMe biobank.

### Germline variant detection and filtering

We implemented two complementary pipelines to discover and jointly genotype germline single-nucleotide variants (SNVs), short insertions/ deletions (indels, and larger structural variants (SVs; ≥50bp). We detected SNVs/indels for each WGS sample GATK HaplotypeCaller before jointly genotyping all SNVs/indels in all samples using the GATK “biggest practices” workflow developed for the Genome Aggregation Database (gnomAD)^18,32^. We subsequently applied variant quality score recalibration (VQSR) filtering to retain only high-quality variants following GATK best practices ^43^.

In parallel, we detected and jointly genotyped germline SVs using our published GATK-SV pipeline ^17^. We executed five independent SV detection algorithms within GATK-SV for each sample: Manta, Wham, MELT, cn.MOPS, and GATK-gCNV using GATK-SV’s recommended parameters ^35,44-48^. After single-sample processing, we divided all samples into six batches of 394 technically-matched samples for multi-sample integration. Following batch specification, we excluded 56 technical outlier samples consistent with standard GATK-SV recommendations ^17^. After initial outlier sample exclusion, we proceeded through GATK-SV multi-sample integration, joint genotyping, and variant resolution using default configurations. After completing the canonical GATK-SV pipeline, we identified and excluded an additional 42 samples that were outliers in the total number of SVs called per genome. We then pruned low-quality non-reference genotypes from our dataset by applying GATK FilterGenotypes, a machine learning model trained to optimize concordance between GATK-SV genotypes with long-read WGS on matching samples from the NIH All of Us Research Program while targeting a maximum false discovery rate (FDR) < 5% ^49^. After pruning low-quality SV genotypes, we outright excluded SV sites that had call rates <95%. We subsequently performed one final layer of outlier sample exclusion, identifying and excluding an additional 60 samples due to persisting as SV call count outliers. Lastly, we performed SV site-level cleanup using a similar procedure to our recent study of germline SVs in pediatric cancer patients ^35^.

### Variant annotation

All germline SNVs and indels were annotated with the Ensembl Variant Effect Predictor (VEP) v110 ^23^, while germline SVs were annotated using GATK SVAnnotate as previously described ^17^. Functional prioritization of rare coding variants was performed using a combination of ClinVar classifications, VEP predicted consequence, and the Rare Exome Variant Ensemble Learner (REVEL) scores, the latter of which predicts pathogenicity of rare missense variants ^24^. Variants were then grouped into one of four functional categories: 1) ClinVar P/LP: variants annotated as pathogenic, likely pathogenic, or conflicting with at least one pathogenic or likely pathogenic entry per the ClinVar release on 02/03/2022, 2) ClinVar P/ LP and/or VEP HIGH impact (frameshift, transcript ablation, splice acceptor, splice donor, stop gained, stop lost, start lost), 3) ClinVar P/LP and/or VEP HIGH impact and/or REVEL score ≥ 0.5, and 4) VEP HIGH or MODERATE (in-frame insertion, in-frame deletion, missense variant, protein altering variant) impact. Variants in these four categories were then annotated with pre-defined gene sets using SnpEff version 5.0c ^50^.

### Genetic ancestry inference

Principal component analysis (PCA) was performed with germline genotypes for all case and control samples using HapMap3 SNPs ^51^. Individuals were then matched into one of five continent ancestry groups defined by the 1000 Genomes Project, including African, American, East Asian, European, or South Asian ^52^. Given the predominance of European ancestry among our cohort, a second PCA was then performed exclusively on the subset of individuals of European ancestry to capture European-specific substructure ^53^. The top five components (PC1–PC5) were used as covariates in downstream analyses to adjust for residual European-specific population stratification.

### Survival analyses

Kaplan-Meier survival analyses and Cox proportional hazards models were performed in the young-onset lung cancer cohort to evaluate associations between overall survival and clinical and molecular features, including sex, stage at diagnosis, oncogenic driver alterations, fusion status, and smoking status. Survival time was defined from date of diagnosis to death or last follow-up. Kaplan-Meier curves were generated using the survival and survminer R packages, with statistical significance assessed using two-sided log-rank tests. Hazard ratios (HRs) and 95% confidence intervals were estimated using Cox proportional hazards models. Models evaluating survival across all stages were adjusted for sex, age at diagnosis, and stage at diagnosis; stage IV–restricted models were adjusted for sex and age at diagnosis, with additional adjustment for histology in sensitivity analyses where specified.

### Polygenic risk scores (PRS)

Polygenic risk scores were calculated in individuals of European ancestry using SNP weights identified in most recent cross-ancestry lung cancer GWAS ^20^. Scores were generated based on 1) top 45 cross-ancestry discovery SNPs associated with lung cancer susceptibility (P < 5×10-8), all SNPs with P < 10-5 in association with lung carcinoma, and 3) all SNPs with P < 10-5 in association with lung adenocarcinoma. PRS was calculated by summing the weighted contributions of each SNP, where weights correspond to the reported effect size of each SNP. A logistic regression model was fit to test association between PRS and case/control status and adjusted for sex and European-specific principle components PC1-5. Raw PRS values were standard-normalized across all samples, and the coefficient for scaled PRS was used to calculate the increase in odds of case status per one-standard-deviation increase in PRS.

### Gene set burden testing

Individual gene sets for gene set enrichment analyses were compiled based on relevant published literature, gene ontology tools, and established cancer pathways. Gene sets with description, references, and component genes are available in Table S4. Separate burden tests for each geneset were performed using only YL cases, only NVS cases, and all cases, with the same controls used for all tests. All tests were restricted to individuals of European ancestry defined above. Additionally, for each geneset and sample set (YL only, NVS only, YL + NVS), four tests were performed using variants from each of the functional consequence categories described above (CV, CV High, CV High & REVEL > 0.5, High & Moderate). Only rare variants, defined here as variants with allele count < 3 in the selected samples, were included in the analysis. Burden tests were implemented as generalized logistic regression adjusted for participant sex and the first 5 European-only PCs used as covariates along with rare variant carrier status for the tested geneset as a binary variable. All resulting p-values were adjusted for multiple testing using the Benjamini-Hochberg FDR procedure ^54^. We defined a gene set as enriched for rare variants if it exhibited FDR q < 0.1; this collection of significant gene sets were used to generate sets of genes for PRS association tests, with one combined gene set per consequence tier. Correlation of combined gene set rare variant burden with both LUAD and lung cancer PRS scores was assessed in young lung cases by Poisson regression with the same covariates used for individual gene set tests for CV, CV High, and CV High & REVEL > 0.5 variants. Regressions using synonymous variant burden in these same genes were performed as a negative control.

### Gene-based rare variant association testing

We performed exome-wide rare variant collapsing burden association tests using SAIGE-GENE+ ^26^ following its recommended procedures. We first created a sparse genetic relatedness matrix using linkage-disequilibrium pruned, common, biallellic SNVs that were also present in the 1000 Genomes Project ^55^. We then used the sparse genetic relatedness matrix to train the SAIGE-GENE+ null model. Covariates included European genetic principal components 1-5 and sex. We tested 18,544 autosomal protein-coding genes annotated in Gencode v47 ^56^ for associations with lung cancer in five scenarios while using all of our controls: all lung cancer cases; young lung cases; never-smoker cases; fusion driven lung cancers; and non-fusion driven lung cancers. Due to subtle subcontinental ancestry differences, likely reflecting differences in cohort recruitment (TOPMED controls: New York City, New York, USA; 54% of never-smoker cases are from Quebec), the never-smoker RVAS analysis demonstrated poor calibration. Therefore, for this analysis only, we excluded European individuals with evidence of admixture from non-European continental ancestries and performed the analysis without principal components included as covariates. For each gene, we further considered four variant impact criteria at AF < 0.1%.

Functional consequence associations were analyzed incrementally, starting with variants annotated in ClinVar as pathogenic or likely pathogenic and sequentially adding variants classifications (i.e., High impact, REVEL > 0.5, and all missense variants). A tradeoff of the SAIGE-GENE+ method when executed with default parameters is that it performs AF-based weighting of variants per gene to boost discovery power; however, due to this variant weighting, the default implementation of SAIGE-GENE+ does not produce interpretable variant effect size estimates. Thus, to enable effect size estimation, we executed SAIGE-GENE+ twice: first to produce maximally powered test statistics to define cancer-associated genes, and a second time while disabling variant reweighting to estimate effect sizes for all genes. Finally, p-values for each pathogenicity were recalibrated using saddlepoint approximation to account for case-control imbalances using a similar implementation as we have previously published ^35,57^.

### Noncoding variant analysis

We evaluated rare noncoding germline variation within regulatory regions linked to a predefined set of candidate genes. For each gene, we defined a cis-regulatory search window extending 500 kb upstream and downstream of the transcription start site (1 Mb range), using GENCODE gene annotations ^56^. Within these windows, candidate regulatory regions were defined using ENCODE/Roadmap epigenomic data from three lung-relevant biosamples: fetal lung (E088), adult lung (E096), and the A549 lung cancer cell line (E114)^58^. For each biosample, regulatory regions were identified by intersecting DNase-seq peaks, representing open chromatin, with H3K27ac ChIP-seq peaks, representing active enhancer and promoter activity. The union of these intersected DNase/H3K27ac regions across the three biosamples was then used to generate a lung-relevant regulatory region set. Germline variants falling within these regions and within the predefined cis-regulatory windows were extracted for downstream association analyses. Rare noncoding variants were defined as variants with minor allele frequency (MAF) <0.001 in the non-Finnish European population of gnomAD ^59^. This approach prioritized noncoding variants located in accessible and transcriptionally active regulatory regions with evidence of activity in lung tissue, fetal lung, or lung cancer cells.

## Supporting information

Supplementary Tables (formatted PDF version)

## Data Availability

All whole-genome sequencing data generated in this study will soon be deposited to the database of Genotypes and Phenotypes (dbGaP).

https://github.com/collins-genomics/young-lung-wgs-pilot

## Acknowledgements

The authors wish to thank the patients who participated in this study and Bruce E. Johnson for careful reading and commenting on the manuscript, Richard Erwin for support in INHERIT study coordination, and Bonnie J. Addario, for a legacy of continued inspiration and support of lung cancer patients and research.

## Funding

JL was funded by LUNGevity Career Development Award. RLC was supported by the Rossy Family Fund at Myriad Canada (formerly KBF Canada) and K99/R00 CA286805 from the US National Cancer Institute. PKB is funded by the Krantz Breakthrough Award, Philanthropic Award from Alexandra Drane, Breast Cancer Research Foundation, National Brain Tumor Society, Terry and Jean de Gunzberg MGH Research Scholar Award, Melanoma Research Alliance, NIH and the Hellenic Women’s Club. MNH was supported by a Research Grant to the institution from Canon Medical Systems, Konica-Minolta, and Addario Lung Cancer Medical Institute.

## Competing Interests

JL is a paid consultant on an unrelated lung cancer study funded by Troper Wojcicki Philanthropies and executed through 23andme. SMY is an employee and shareholder of Labcorp. BJG is an employee of Roche/Genentech and holds stock in Roche. AAA receives research funding from Varian and NH TherAguix/TheraGuix. PKB has consulted for Voyager Therapeutics, Tesaro, SK Life Science, Sintetica, Pfizer, Merck, ElevateBio, Dantari, Angiochem, MPM, Medscape, Exelixis, Kazia, InCephalo, Genentech, Eli Lilly, CraniUS, Axiom, Atavistik, and Advise Connect Inspire, serves on the Scientific Advisory Board for Selectin Therapeutics (with equity), Kazia, and CraniUS, and has received Speaker’s Honoraria from Genentech and Pfizer. She has received institutional research support (to MGH) from Kinnate, Mirati, Merck, and Eli Lilly and clinical trial support (to MGH) from AstraZeneca, GSK, Pfizer, Merck, Genentech-Roche, Bristol Myers Squibb, Mirati and Kazia. JJN is a paid consultant for Aadi Biosciences, ANP Technologies, AstraZeneca, BioAtla, Bristol Myers Squibb, Catalym, Daiichi Sankyo, Gilead, Genentech, Kalivir, Merus, Sanofi, Pfizer, Takeda, Tubulis, and Urogen; receives research funding from Genentech and Merck; holds intellectual property with Cansera; and holds ownership interests in Affyimmune, Cansera, Epic Sciences, Indee Bio, Quantgene, Amgen, Johnson & Johnson, and Novartis. NF reports advisory and consulting relationships with Johnson & Johnson, Sanofi, Pfizer, Mirati, Bristol Myers Squibb, Merck, Regeneron, Takeda, Lilly, Daiichi Sankyo, EMD Serono, Novartis, Novocure, Jazz Pharmaceuticals, Genentech-Roche, Nuvalent, Nuvation Bio, Catalyst Pharmaceuticals, Caris, and AstraZeneca. She has received research funding from the American Society of Clinical Oncology (ASCO), the International Association for the Study of Lung Cancer (IASLC), the Lungevity Foundation, the Harvard Cancer Center Lung Cancer SPORE, the Champa Fund for Early Cancer Detection, the GO2 Foundation for Lung Cancer, the Women’s Health Access Matters (WHAM) Foundation, Daiichi Sankyo, Genentech, AstraZeneca, Johnson & Johnson, Nuvation Bio, and Pfizer. She has also participated in speaking engagements for Clinical Care Options (CCO), Dava Oncology, Ideology Health, Oncology Central, CME Outfitters, ASCO Post, OncLive, and Physician Education Resource (PER). EMV reports advisory and consulting roles with the Novartis Institute for Biomedical Research, Serinus Bio, TracerBio, and Cellyrix. He has received research support from Novartis and Bristol Myers Squibb (BMS). He also reports equity interests in Tango Therapeutics, Enara Bio, Manifold Bio, Microsoft, Monte Rosa, Serinus Bio, TracerBio, and Cellyrix. In addition, he is an inventor on institutional patents related to chromatin mutations and immunotherapy response, as well as methods for clinical interpretation, and has participated in intermittent legal consulting on patents for Foaley & Hoag. PAJ is a paid consultant for AstraZeneca, Boehringer Ingelheim, Pfizer,Roche/Genentech, Chugai Pharmaceuticals, Eli Lilly pharmaceuticals, SFJ Pharmaceuticals, Voronoi, Daiichi Sankyo, Biocartis, Novartis, Sanofi, Takeda Oncology, Mirati Therapeutics, Transcenta, Silicon Therapeutics, Syndax, Nuvalent, Bayer, Eisai, Allorion Therapeutics, Accutar Biotech, Abbvie, Monte Rosa Therapeutics, Scorpion Therapeutics, Merus, Frontier Medicines, Hongyun Biotechnology, Duality Biologics, Blueprint Medicines, Dizal Pharma, GlaxoSmithKline, Tolremo, Myris Therapeutics, Bristol Myers Squibb; receives research funding through DFCI from AstraZenenca, Daiichi Sankyo, PUMA, Eli Lilly pharmaceuticals, Boehringer Ingelheim, Revolution Medicines, Takeda Oncology, and Troper Wojcicki Foundation; receives post-marketing royalties from Dana Farber Cancer Institute owned intellectual property on *EGFR* mutations licensed to Labcorp. The remaining authors declare no competing interests.

## Data Availability

Whole-genome sequencing data generated in this study will be deposited in the database of Genotypes of Phenotypes (dbGaP) prior to publication, available under controlled access because of participant privacy considerations. Summary statistics from gene-set burden analyses, exome-wide association analyses, structural variant burden analyses, and polygenic risk score analyses are provided in the Supplementary Tables. Publicly available datasets used in this study include the Sherlock-Lung cohort (phs001697.v1.p1), BioMe (phs001644.v3.p2), AACR Project GENIE (version 19.0), gnomAD v4.1, and TCGA.

## Code Availability

All custom code used for data processing, quality control, statistical analyses, and figure generation is available at https://github.com/collins-genomics/young-lung-wgs-pilot. Analyses were performed using publicly available software packages including GATK-HC, GATK-SV, REGENIE, PLINK, bcftools, and R packages as described in the Methods.

## Notes

### Author Declarations

The Dana-Farber Cancer Institute institutional review board gave ethical approval for this work. Secondary genomic analyses conducted for this study were approved under DFCI-IRB protocols 02-180, 14-183, and 18-411.

