## Supplementary Tables (formatted PDF version) for "Germline determinants of risk and molecular subtype in young-onset lung cancer"

### Supplementary Information for LoPiccolo* & Collins* *et al.*

### SUPPLEMENTARY TABLES

1. Associations Between Age at Diagnosis and Somatic Driver Alterations in Lung Adenocarcinoma (LUAD) from AACR Project GENIE
2. Distribution of Somatic Driver Alterations in Young-Onset Lung Adenocarcinoma (LUAD) and TCGA Comparison Cohorts.
3. † Clinical, Demographic, and Sequencing Characteristics of Study Population
4. *Curated Gene Sets and Pathways Evaluated for Rare Germline Variant Burden Analyses
5. *Gene-Set Burden Analysis of Rare Coding Germline Variants in Young-Onset and Never-Smoking Lung Cancer
6. *Association Between Polygenic Risk Scores and Rare Germline Variant Burden in Young-Onset Lung Cancer Cases
7. *Exome-Wide Gene-Based Rare Variant Association Results in Young-Onset and Never-Smoking Lung Cancer
8. *Gene-Set Burden Analysis of Rare Germline Structural Variants in Young-Onset and Never-Smoking Lung Cancer
9. *Association Testing of Rare Noncoding Germline Variants in Candidate Regulatory Elements

† To be deposited on dbGaP upon publication
* Accessible as downloadable files

| Gene Name | OR | 95% CI (Lower) | 95% CI (Upper) | P-Value |
|---|---|---|---|---|
| *EGFR* | 1.201449117 | 1.017767329 | 1.413976032 | 0.028709434 |
| *KRAS* | 0.315185562 | 0.25365892 | 0.388391907 | 4.30E-35 |
| *ALK** | 8.321568904 | 6.391459979 | 10.74384401 | 1.41E-42 |
| *ROS1** | 5.887259231 | 3.890669216 | 8.679367949 | 3.97E-14 |
| *RET** | 3.553850585 | 2.157789808 | 5.593932657 | 1.60E-06 |
| *MET* | 0.144004269 | 0.029564897 | 0.424621465 | 4.23E-06 |
| *ERBB2* | 1.876823551 | 1.259797602 | 2.709095228 | 0.001446076 |
| *BRAF* | 0.364949235 | 0.156254762 | 0.727517692 | 0.001516454 |
| *NTRK1** | 7.859711336 | 1.879262949 | 25.09458431 | 0.003150302 |
| All Fusions* | 7.324860383 | 5.981638089 | 8.926264337 | 2.34E-64 |
| Other/WT | 1.138264073 | 0.979316647 | 1.321283014 | 0.086292266 |

**A. Associations between oncogenic driver alterations and age at diagnosis in LUAD**

| Gene Name | Coefficient | Standard Error | P-Value | Increase Per Year (%) |
|---|---|---|---|---|
| *EGFR* | -0.008247734 | 0.001441664 | 1.06E-08 | -0.82% |
| *KRAS* | 0.014914782 | 0.001351873 | 2.66E-28 | 1.50% |
| *ALK** | -0.067756945 | 0.003970638 | 2.73E-65 | -6.55% |
| *ROS1** | -0.059129184 | 0.005594667 | 4.16E-26 | -5.74% |
| *RET** | -0.045424847 | 0.00577319 | 3.60E-15 | -4.44% |
| *MET* | 0.067925928 | 0.004708143 | 3.48E-47 | 7.03% |
| *ERBB2* | -0.019839821 | 0.003913764 | 3.99E-07 | -1.96% |
| *BRAF* | 0.010891399 | 0.003933758 | 0.005628029 | 1.10% |
| *NTRK1** | -0.068824205 | 0.017090202 | 5.65E-05 | -6.65% |
| All Fusions* | -0.064282604 | 0.002941209 | 6.84E-106 | -6.23% |
| Other/WT | -0.00139538 | 0.001293659 | 0.280752717 | -0.14% |

**B. Linear regression analysis of age-associated enrichment of oncogenic driver alterations**

**Supplementary Table 1. Associations Between Age at Diagnosis and Somatic Driver Alterations in Lung Adenocarcinoma (LUAD) from AACR Project GENIE.**

* denotes genes somatically implicated in oncogenic fusions.

| Driver Gene | Alteration | Young Lung Count (%) | TCGA >45 Count (%) |
|---|---|---|---|
| *EGFR* | Exon 19 deletion | 62 (69.7%) | 17 (34%) |
| | L858R | 12 (13.5%) | 21 (42%) |
| | Exon 20 insertion | 7 (7.9%) | 2 (4%) |
| | Exon 18 deletion | 0 | 3 (6%) |
| | G719X/S768X | 4 (4.5%) | 3 (6%) |
| | L861Q | 0 | 3 (6%) |
| | Exon 18-25 duplication | 1 (1.1%) | 0 |
| | Other | 1 (1.1%) | 1 (2%) |
| | Fusion | 2 (2.3%) | 0 |
| *KRAS* | G12A | 1 (4.4%) | 14 (9.7%) |
| | G12C | 7 (30.4%) | 64 (44.4%) |
| | G12D | 13 (56.5%) | 16 (11.1%) |
| | G12S | 0 | 4 (2.8%) |
| | G12V | 1 (4.5%) | 31 (21.5%) |
| | G13C/D | 0 | 7 (4.9%) |
| | Q61X | 1 (4.5%) | 3 (2.1%) |
| | Other | 0 | 5 (3.5%) |
| *HER2* | Exon 20 insertion | 9 (69.2%) | 4 (100%) |
| | V659E | 2 (15.4%) | 0 |
| | G660D | 1 (7.7%) | 0 |
| | L869R | 1 (6.7%) | 0 |
| *BRAF* | V600E | 1 (50%) | 7 (25%) |
| | Non-V600E | 1 (50%) | 21 (75%) |
| *MET* | Exon 14 | 0 | 6 (85.7%) |
| | Amplification | 2 (100%) | 0 |
| | Fusion | 0 | 1 (14.3%) |

**Supplementary Table 2. Distribution of Somatic Driver Alterations in Young-Onset Lung Adenocarcinoma (LUAD) and TCGA Comparison Cohorts.**

Percentages represent the proportion of each alteration among all alterations within that driver gene in the respective cohort.
